# Individual patterns of activity predict the response to physical exercise as an intervention in mild to moderate depression

**DOI:** 10.1101/2024.02.12.24302721

**Authors:** Stefan Spulber, Sandra Ceccatelli, Yvonne Forsell

**Affiliations:** Department of Neuroscience; Department of Global Public Health, Karolinska Institutet, 171 77 Stockholm, Sweden

## Abstract

Physical exercise (PE) as antidepressive intervention is a promising alternative, as shown by multiple meta-analyses. However, there is no consensus regarding optimal intensity and duration of exercise, and there are no objective criteria available for personalized indication of treatment. The aims of this study were (1) to evaluate whether individual activity patterns before intervention can predict the response to treatment; and (2) to evaluate whether the patient outcome can be improved by using prior information on treatment efficacy at individual level. The study included subjects with mild to moderate depression randomized to three levels of exercise intensity in the Regassa study. Using a previously developed pipeline for data analysis, we have generated linear regression ensembles to predict the response to treatment using features extracted from actigraphy recordings. To understand the contribution of individual features, we performed a Bayesian analysis of coefficients, and found that different levels of PE intensity yield distinct signatures in enriched feature subsets. Next, we used the trained ensembles for a counterfactual analysis of response and remission rate provided prior knowledge of response to treatment outcome. The response to either PE regime was estimated for all patients, irrespective of original treatment assignment. Each patient was then virtually allocated to the PE regime predicted to yield best outcome, and the response and remission rates were compared against simulated random assignment to treatment. The counterfactual analysis showed that assignment to best individual PE regime yields significantly higher (increase by 28%) remission rates as compared to random assignment to treatment, which is accounted for by improved response in about 32% of the patients as compared to observed treatment outcome. While it is not possible to claim individual protocol optimization, our data suggest it may be possible to identify a PE regime to yield the best results for a patient based on individual circadian patterns of activity.

## Introduction

The world-wide prevalence of depression is reported to be 4.4% and it is increasing, especially in young persons [1]. Depression is a recurrent often life-long disorder and long-term recovery is low, approximately 60% at 2 years, 40% at 4 years, and 30% at 6 years [2].

Due to the large number affected and the chronicity there is a need for treatment options that are easily accessible, have minor side effects and are cost-effective. Today the treatments that are available is mainly different types of psychotherapy and antidepressant medication. However, psychotherapy is costly and not always available, while antidepressant medications have side-effects. The adherence to antidepressants is quite low, at a six-month examination approximately 50% had discontinued the treatment [3]. Additionally, even intensively monitored antidepressant treatment have low response rates; the STAR*D trial looked at remission rates after level 1 to 4 treatments and found a remission rate of 35% [4]. Response rates after psychotherapy are almost equally low, a meta-analysis reported 46.3% in mild to moderate depression [5] There is also a large treatment gap, only one-third of those affected receive adequate treatment [6].

Physical exercise (PE) is a promising therapy, since there is evidence from multiple meta-analyses showing good effects on depression [7–9]. There are no side-effects, on the contrary physical exercise has other positive effects on overall somatic health. Depression is considered as a risk factor for cardiovascular disorders and thus the potential positive benefit of exercise might be of even greater value [10]. Despite mounting evidence, PE is rarely used in a structured way. Adherence is a common problem for antidepressive therapies, and exercise is no exception. A large study reported that 50% of non-depressed individuals who start to exercise do not continue after six months [11]. Due to the nature of depression with symptoms like lack of energy and initiative, adherence is probably even lower in depressed individuals. It is problematic to look at adherence in clinical studies of exercise as a treatment since those involve extensive support that most likely is not feasible to use in daily clinical practice. Currently there is no consensus regarding optimal intensity and duration of exercise as antidepressive treatment, and there are no objective criteria available for personalized indication of treatment. The aims of this study were (1) to evaluate whether individual activity patterns before intervention can predict the response to treatment; and (2) to evaluate whether the patient outcome can be improved by using prior information on treatment efficacy at individual level. The study included subjects with mild to moderate depression randomized to three distinct PE regimes, which allowed a counterfactual analysis of potential improvement in patient outcome when treatment allocation uses *a priori* information on predicted response.

## Materials and Methods

### Subjects

Data derived from the Regassa study, a single blind, multi-center randomized controlled trial conducted in six Swedish regions (Stockholm, Kronoberg, Blekinge, Skåne, Västra Götaland and Västmanland) (for further description see [12]). The main aim was to compare treatment as usual, physical exercise, and Internet-based cognitive behavioural therapy as treatments for mild to moderate depression in primary care patients. The study was registered with the German Clinical Trial Register (DRKS study ID: DRKS00008745) and approved by the Stockholm regional ethical review board (Dnr: 2010/1779-31/4). Inclusion criteria were primary care patients, aged 18–67 years with mild to moderate depression according to Montgomery-Åsberg Depression Rating Scale (MADRS) [13]. Exclusion criteria were a primary diagnosis of alcohol or drug dependency, serious somatic disorder, or requiring specialist psychiatric treatment. Subjects for this study were recruited between 2011-02-04 and 2014-02-28. All subjects provided informed consent for participation in the study after having received written information about the study. This study is based on reanalysis of data collected for the Regassa study, and for this purpose, the data was accessed starting on 2018-05-07. The authors did not use any information that could identify individual participants and used exclusively anonymized records.

### Physical exercise intervention and measurements

Before starting and the week after the participants ended the 12-weeks intervention, they wore a triaxial accelerometers (ActiGraph GT3X+, ActiGraph LLC, Pensacola, FL, USA) to measure physical activity level. The instructions were to wear the accelerometer for seven days on the right hip, continuously throughout the waking part of the day, removing it only for bathing or swimming.

The subjects randomized to physical activity were further randomized to one of three exercise intensity levels as follows: (1) light exercise, *e.g.*, flexibility and balance exercises; (2) moderate exercise-intermediate-level aerobics; and (3) vigorous exercise-more strenuous aerobics. Participants were asked to attend three classes of 55 min duration per week for 12 consecutive weeks. The classes were offered at a modern fitness centre organization with premises throughout Sweden and all participants received free membership.

To ensure that the intensity levels differed between the recommended gym classes, members of the research team designed classes of differing intensity that were tested using indirect calorimetry and pulse watches in a laboratory setting. They then attended different classes at the gym using pulse watches and compared the results with the laboratory classes. The results were used to select classes with different intensity levels to recommend to the participants.

To see if the participants exercised at the assigned intensity level, they wore pulse watches during the exercise classes. These data were collected each week. The results showed that the three groups differed from each other on the average proportion of calculated maximal heart rate and minutes spent above 80% of maximal heart rate (MHR) [14]. The primary outcome of this study was the change in severity of depression, assessed as percentage change in MADRS after treatment.

### Data processing and feature extraction

The workflow is depicted in Fig. 1. Out of 316 patients assigned to PE as antidepressant intervention, 168 had actigraphy recorded. The raw data was exported using the proprietary software and processed in Matlab® (version 9.x; The MathWorks Inc, Natick, MD, USA) using custom scripts (see also [15,16]). The quality control was performed by the same observer, blind to group belonging. All recordings were first inspected visually using a standardized procedure designed to identify stretches of missing data, artifacts, and gross abnormal circadian patterns of activity (*e.g.*, shift work, or other consistent activity at night). We considered a maximum of 8 consecutive days at the very beginning of the recording period (*i.e.,* before starting PE as antidepressant intervention). Recordings with less than 3 consecutive days within the first week were excluded. This procedure yielded usable recordings for 100 patients. For further analyses, we used only recordings from patients which were not taking antidepressant medication at the time of recording actigraphy data (66 patients, in total 383 recorded days; Fig. 2A). All recordings were cropped between first and last recorded midnights to restrict the length to an integer number of 24-h periods, then the onset and offset of activity recording for each recorded day was identified as the boundaries of the longest stretch of inactivity during device recharging sessions (Fig. 2A). The complete list of features extracted is available in Supplementary Table 1. Where possible, the features were calculated for each recorded day, or on 3 consecutive days with wrap-around for first and last days recorded and used for estimation of variability. In addition, a complete set of features was calculated on the full-length recording. The following features were measured: circadian period; circadian peak; scaling exponents [17]; intradaily variability (IV); interdaily stability (IS) [18]; day-to-day variability; and propensity to sustain activity. The subjects were instructed to recharge the actigraphs overnight, which limited the recording to the active phase exclusively. This provided an approximation of activity onset and offset but did not allow the estimation of activity during daily trough (presumably reached during resting period) or relative amplitude of circadian rhythms [19,20]. Circadian period was estimated using the Lomb-Scargle algorithm implementation optimized for Matlab [21]. The Lomb-Scargle periodogram was preferred over the commonly used Sokolove-Bushell algorithm [22] because the latter has been shown to yield period estimates biased towards periods below 24 h [23]. The circadian period was calculated over the entire recording using an oversampling factor of 10 to yield a resolution in the range of minutes for the estimate. The scaling exponent for detrended fluctuation analysis was calculated for the magnitude of measured activity in 1-min bins using boxes equally spaced on a logarithmic scale between 4 min (4 consecutive samples) and 24 h (1440 consecutive samples) as described by Hu et al. [17]. The scaling exponent is a feature of the intrinsic regulatory mechanisms controlling the rest/activity patterns. It has not been shown to be sensitive to extrinsic factors the subject is exposed to in normal daily activity but is altered because of disease [17,24,25]. Intradaily variability (IV) estimates the fragmentation of activity patterns by calculating the ratio between mean squared differences between consecutive time intervals and the mean squared difference from global mean activity per interval; it increases as the frequency and the magnitude of transitions between rest and active intervals increase and decreases as the active and inactive intervals consolidate [26]. Interdaily stability (IS) evaluates the coupling between activity patterns and circadian entrainers and is calculated as the ratio between variability around circadian profile and global variability. High values indicate consistent activity patterns across days, consistent with strong coupling between activity and circadian entrainers. The day-to-day variability comprised 3 features as follows: circadian profile variance between consecutive days (rmssd), calculated as Euclidean distance between consecutive days, normalized to the total number of samples per day; variation from average circadian profile rmsep), calculated as the Euclidean distance between each day and the average profile, normalized to the total number of samples per day; and the normalized difference between consecutive days (ddv), calculated as the ratio between mean difference between circadian profiles of consecutive days and mean deviation from average circadian profile. The propensity to sustain activity (pHiSlope) was calculated as the slope of likelihood to sustain or increase activity in the next minute against current level of activity. The distribution was calculated for minutes with activity count>10 (which eliminated the range specific for sleep, ∼30% of datapoints/day) in 20 equally spaced bins covering the range up to the 99^th^ percentile of active minutes. This is a hybrid measure applying a sequence-based analysis on the distribution of activity counts/min (assumed to have exponential distribution). The likelihood to further increase activity drops with increasing the activity counts in the current minute, therefore the slope is negative, and approaches 0 at the right tail of the distribution. A flat slope indicates the subject is unlikely (possibly not willing) to sustain even low levels of activity. For scrambled timeseries (preserved distribution, but random sequence), the slope is about -0.3 (likelihood to sustain activity decreases by 30% when activity increases 10 -fold).

**Figure 1.**
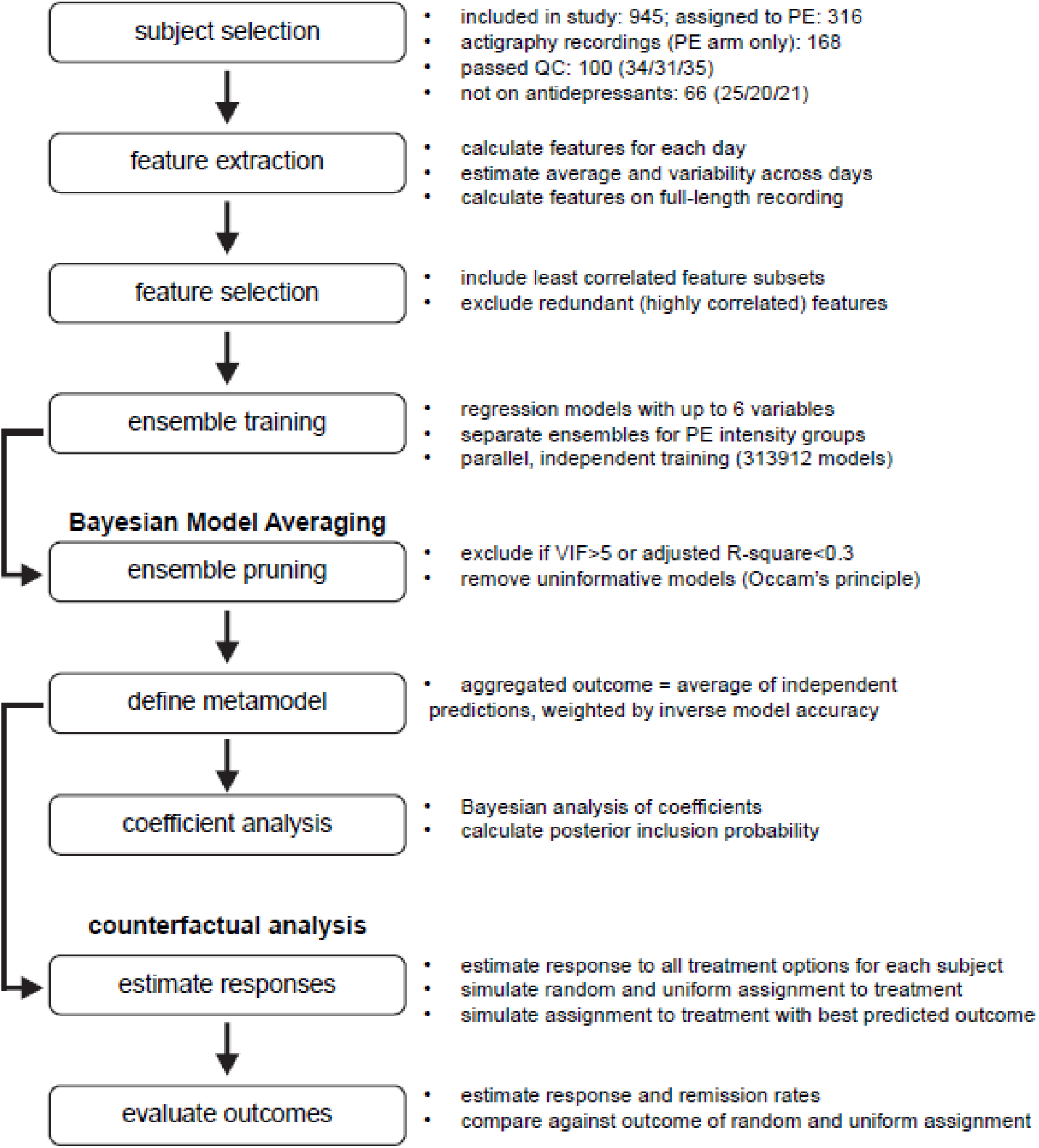
Graphic description of workflow for data analysis.

**Figure 2.**
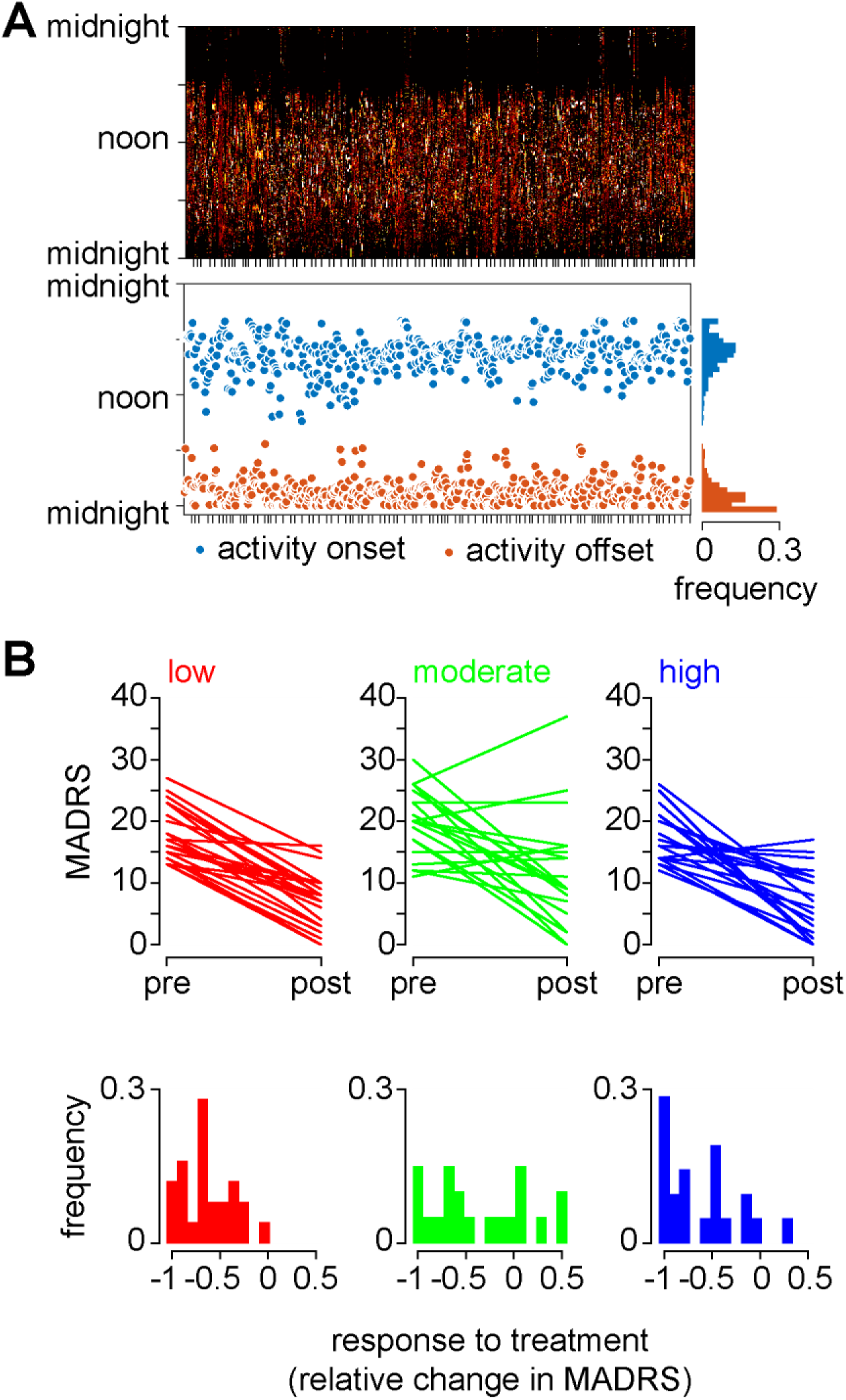
Description of data used for model training. (A) Top panel: heatmap of normalized activity used for feature extraction. Individual days recorded form each subject shown in temporal order of recruitment, independent from treatment assignment. Bottom panel: location of activity onset and offset for each day (corresponding columns in the top panel). Ticks on horizontal axis indicate the end of the recording for each patient. (B) Change in MADRS score recorded for each patient assigned to specific PE regime.

### Feature selection

There is virtually no consensus around parameter selection for actigraphy. To mitigate the impact of multicollinearity, we used a hybrid heuristic and data driven feature selection procedure to facilitate the interpretation of results from Bayesian model averaging. First, we investigated the correlations between daily averages, sequential variability, variability around average profile, and full-length recording, and selected the sets with least significant correlations for further analyses: variability around average profile and full-length recording (Supplementary Fig. S1). The features included displayed various degrees of similarity, as illustrated by the matrix of correlations in Supplementary Fig. S2. Starting from the observation of clusters of conceptually related and highly correlated features, we reduced the number of features to be used for ensemble training as follows: from each cluster we kept only one feature (the one with lowest number of correlations overall), including both full-length recording and daily variability. The feature space consisted of 26 features (Supplementary Fig. S2).

### Ensemble training and Bayesian model averaging

We modelled the response to treatment, estimated as *(MADRS_post_-MADRS_pre_)/MADRS_pre_*using ensembles of multivariate linear regressions with features describing activity patterns as predictors. The changes in MADRS in patients included in model training procedures is shown in Fig. 2B. Three independent ensembles were generated -one for each PE regime. Each initial ensemble was generated by independent homogenous training using a systematic bootstrapping scheme (with replacement) for selecting up to 6 features/model, which yielded 313 912 models/ensemble. The pruning procedure consisted of an application of Occam’s window algorithm [27,28]. First, we excluded all models satisfying the following criteria: VIF>5 for any single feature in the model (to avoid multicollinearity); or adjusted R-square<0.3. Next, we excluded all models receiving less support from the data than their simpler sub-models. Thus, more complex models (using *n* features) were excluded if they are less accurate than any lower complexity models trained on the same set of features (using any combination of *k<n* subsets). This procedure penalizes more complex models if their accuracy is not superior to simpler models. Lastly, we defined the metamodel (aggregated ensemble output) as the mean of predictions from individual models weighted by the inverse of model RMSE [27]. To evaluate the performance of each ensemble, the models were sorted by increasing RMSE (decreasing accuracy) and the metamodel performance was evaluated in a cumulative fashion. The aggregated output was then used for estimating the precision and accuracy, estimated by adjusted R-square and RMSE, respectively.

### Bayesian coefficient analysis

The prior inclusion probability for any individual feature was calculated as the number of models including a specific feature relative divided by the total number of models possible to train under the constraint of maximum 6 features/model (see Supplementary Material for calculation), which yielded a value of 0.2179. The posterior inclusion probability (PIP) for each feature was calculated as the frequency of occurrence in the pruned ensembles, and was used for defining levels of evidence strength as follows: PIP>0.2179 identifies features with frequency of occurrence increased as compared to initial probability (enriched), and indicates medium to strong evidence of correlation; in contrast, PIP < prior inclusion probability denotes features depleted after pruning, and indicates weak evidence of correlation. The effect size for individual features was estimated as the average of standardized coefficients across all models in the pruned ensemble. The stability of individual features (context-independence) was evaluated using the coefficient of variation (CV=standard deviation/average) for standardized coefficients.

### Counterfactual analysis

The purpose of this calculation was to estimate the potential impact of prior knowledge on response to treatment on patient outcome. We used the pruned ensembles to estimate the response to either treatment for each patient, independent from treatment allocation in the original clinical trial. The predicted responses were then filtered to remove predictions below -1 (*i.e.*, yielding negative values for MADRS after treatment), and kept the best predicted response for each patient. The symptom severity after treatment (*MADRS_post_*) was estimated as *MADRS_post_* = [(1 + *response*)*MADRS_pre_*], where *MADRS_pre_* is the symptom severity score measured before treatment, and *response* is the best estimated response to any of the treatment options, and the result was rounded to nearest integer. The response rate was calculated as the proportion of patients in which the MADRS score decreased by more than 50% of the score measured before intervention. The threshold for remission was set at 10, and the remission rate was calculated as the portion of patients with MADRS<10 after intervention.

## Results

A description of the study sample is presented in Table 1. The subjects with complete accelerometer data had a lower MADRS mean score and were less often unemployed than those that did not provide complete accelerometer data. The feature selection procedure yielded only sparse differences across groups or genders within the population used for modelling the response to treatment (Supplementary Fig. S2).

**Table 1.** Description of the participants randomized to physical activity with and without complete accelerometer data.

| Characteristics | All n=316 | With complete accelerometer data n=66 |
| --- | --- | --- |
| Gender |  |  |
| Male, n(%) | 74(23.4) | 21(31.8) |
| Female, n(%) | 176(55.7) | 45(68.2) |
| Mean age (SD) | 42.3(12.2) | 42.6 (11.9) |
| Exercise group |  |  |
| Light,n(%) | 106(33.5) | 25(37.9) |
| Moderate,n(%) | 106(33.5) | 20(30.3) |
| Vigorous,n(%) | 104(32.9) | 21(31.8) |
| Have a tertiary education,n(%) | **128**(40.5) | **26**(39.4) |
| Born outside Sweden,n(%) | 67(21.2) | 15(22.7) |
| Unemployed,n(%) | **37**(11.7) | **2\***(3.0) |
| MADRSpre, mean(SD) | 22.2(6.9) | 20.3(5.1)* |
| MADRSpost, mean(SD) | 11.4(7.9) | 10.2(7.3) |
| Response (relative change in MADRS), mean(SD) | -0.43(0.54) | -0.48(0.35) |

\*p<0.01

### Aggregated prediction performance

First, we asked whether individual features describing the patterns of activity before treatment correlate with the magnitude of response to treatment, and whether the pattern of correlations is treatment specific. We found distinct patterns of correlations with the response to treatment (Fig. 3A), and the correlations surviving Benjamini-Hochberg FDR correction can be summarized as follows: better response to low intensity PE correlates with more variable scaling exponent; better response to moderate intensity PE correlates with lower amplitude of circadian peak and lower variability of circadian profile; and better response to high intensity PE correlates with stronger circadian entrainment. The maximum coefficient of determination (R-square) for individual features was 0.19, 0.16 and 0.22 (corresponding RMSE: 0.22, 0.42, and 0.31) for low, moderate, and high intensity physical activity, respectively.

**Figure 3.**
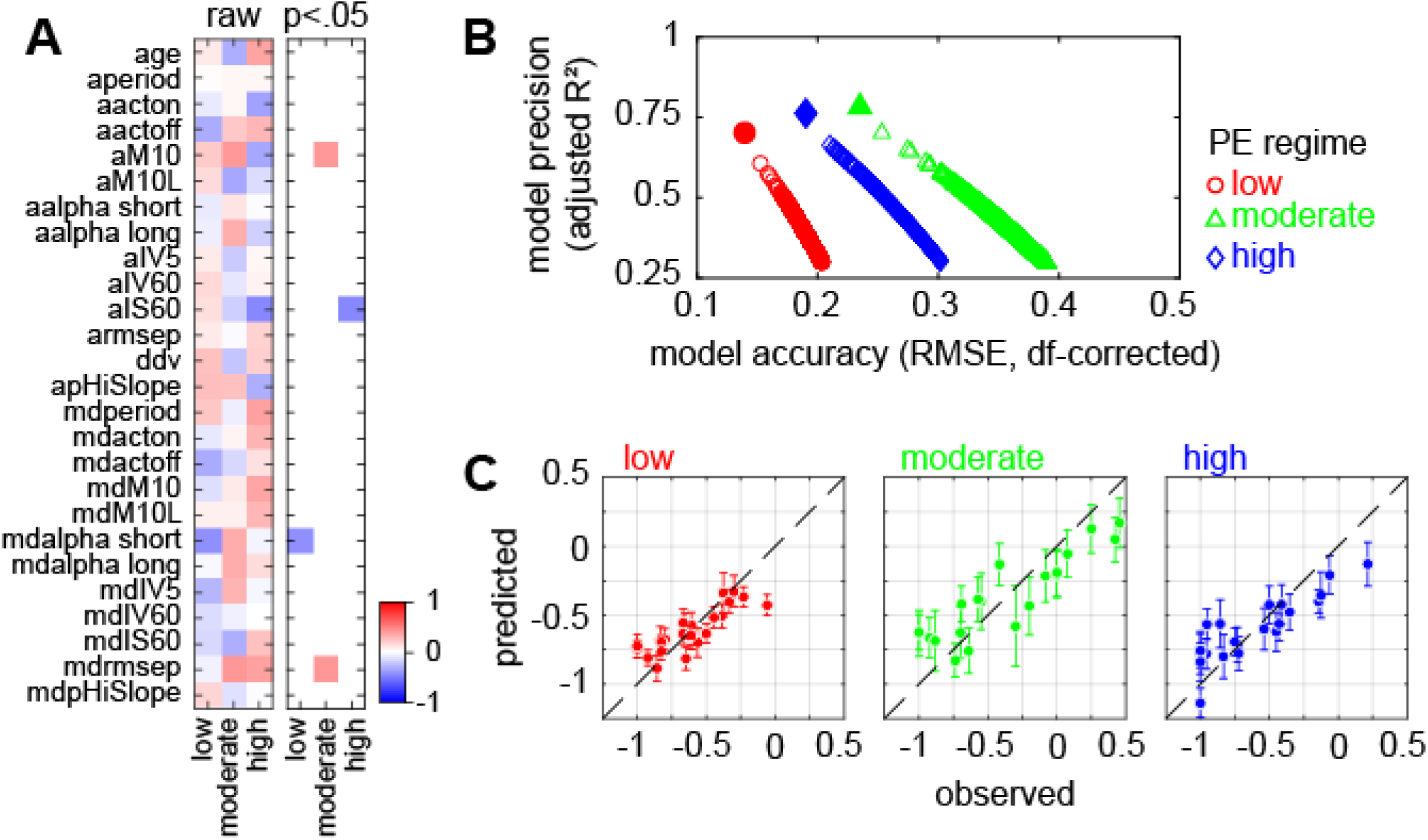
Correlations with response to treatment. (A) Correlations between individual features and response to treatment. Note the distinct patterns of direction and magnitude of linear correlations across groups. Significant correlations displayed in the right panel. No correlation survives FDR correction for multiple comparisons. (B) Comparison between aggregated prediction (filled symbols) and performance of individual models trained to fit the response to treatment (empty symbols). Note that the precision and accuracy of aggregated ensemble prediction is superior to any individual model. (C) Calibration plots for ensemble prediction of response to PE as antidepressant intervention. Corresponding performance measures are shown in (B) as filled symbols. Error bars depict weighted standard deviation of metamodel prediction error.

Next, we trained linear regression ensembles using a parallel, independent training procedure, followed by model pruning. This yielded ensembles of multivariate linear regression models widely outperforming individual single features. The pruned ensembles consisted of 3503, 2088, and 6423 models with coefficient of determination >0.3 for low, moderate, and high intensity, respectively (Fig. 3B, see also Supplementary Fig. S3). We evaluated the aggregated accuracy as the cumulative average of predicted individual responses, weighted by the inverse model RMSE for each treatment option (Fig. 3C). The aggregated accuracy displayed distinct minima, followed by progressive degradation as the accuracy of included models increased. The best accuracy achieved was 0.12 (N=13 models), 0.15 (N=126 models) and 0.15 (N=17 models) for low, moderate, and high intensity physical activity, respectively, which corresponds to an increase in aggregated accuracy between 1.8 and 2.6 times as compared to single models.

### Analysis of coefficients and feature enrichment

The importance of the variables in a model can be assessed using various approaches [29]. In an ensemble training procedure, however, the contribution of individual features may change depending on the context, i.e., the subset of features included in the model. We assessed the importance of features by means of a Bayesian analysis of coefficients and used the following parameters to evaluate the contribution of individual features: PIP (to estimate how relevant a specific feature is across all models in an ensemble); mean standardized coefficient (to estimate effect magnitude across all models); and coefficient of variance (to estimate the context-dependence) (Fig. 4A, Supplementary Fig. S4). These three measurements are not necessarily orthogonal, but their joint analysis highlights the most relevant features and supports the interpretation of correlation coefficients. Of note, the actual values of these parameters change with the size of the ensemble (*i.e.*, they depend on how restrictive the pruning criteria are), but the overall ranking by PIP is rather stable, and the standardized coefficients change only marginally (Fig. 4A: all enriched features have CV<0.6).

**Figure 4.**
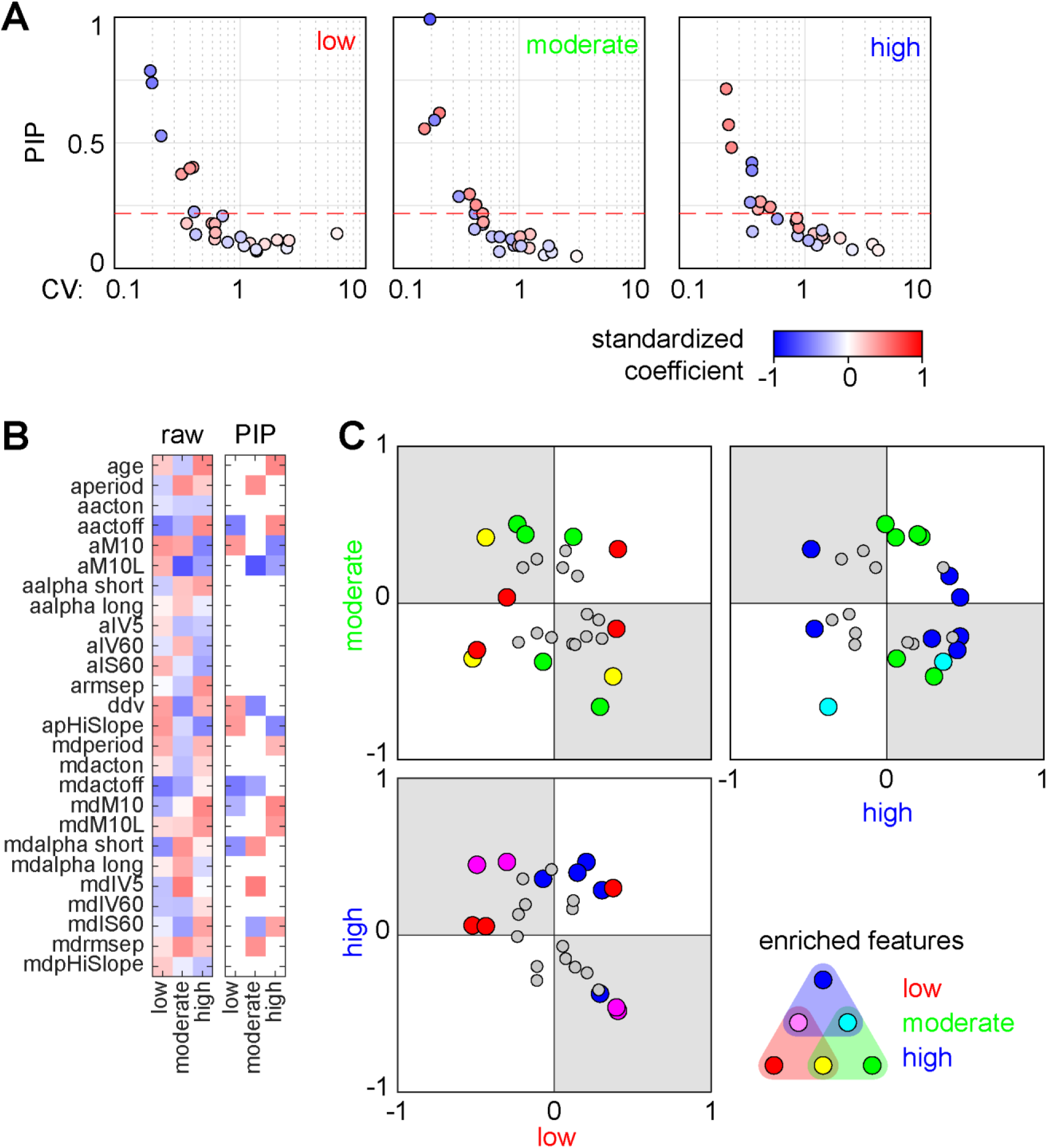
Bayesian analysis of coefficients. (A) Synoptic view of effect size (color-coded), variability (CV), and probability of inclusion in models (PIP) for individual features. Features with PIP higher than the initial inclusion probability (0.2179, indicated by horizontal red dashed line) are defined as enriched. (B) Illustration of differences in ensemble composition. The heatmap in the first panel shows average standardized coefficients. The second panel display only the coefficients with PIP > 0.2179 (see (A) for reference). Note the distinct signatures for each ensemble. (C) Direct comparison between effect sizes of individual features across ensembles. Enriched features highlighted and color-coded based on differential enrichment in each ensemble. Features mapping in top-right and bottom-left quadrants indicate similar effects, while features mapping in top-left and bottom-right quadrants (shaded) indicate opposite effects between ensembles.

We next focused on the subset of enriched features, particularly asking whether individual features have similar contributions across ensembles (Fig. 4B). The subsets of enriched features expanded beyond the subset of features displaying significant correlation with the response to treatment for low intensity physical activity, while for moderate intensity there was only partial overlap (1 out of 2 features with significant correlation with the response was enriched in the final ensemble), and no overlap for high intensity physical activity (compare Fig. 3A and Fig. 4B). The subsets of features enriched yielded distinct signatures for each ensemble, with 9 out of 16 enriched features shared between ensembles (Fig. 4B). The subsets of enriched features can be summarized as follows: for low intensity PE, better response is predicted by later and more variable offset of activity, lower, and less variable circadian peak of activity, lower variability of circadian profile and higher propensity to sustain activity; for moderate intensity PE, better response is predicted by weaker circadian entrainment, lower variability of circadian profile, and later circadian peak of activity ( -aM10L); and for high intensity PE, better response is predicted by younger age, a circadian peak of activity which is high, late, and less variable across days, overall stronger circadian entrainment, and lower propensity to sustain activity. To better visualize the differential impact of individual features across ensembles, we plotted the mean standardized coefficient in 2D (Fig. 4C), and found only 1 out of 9 shared features between pairs of ensembles to have similar impact in both ensembles. The distribution of standardized coefficients across the quadrants indicates that (1) the prediction of response to moderate intensity PE is to a large extent independent from both low and high intensity PE; and (2) predicted response to low intensity PE is inversely correlated with the predicted response to high intensity PE (Supplementary Fig. S5).

### Improvement in patient outcome provided a priori knowledge of individual response to treatment

We designed a counterfactual analysis assuming *a priori* information on individual response to different PE regimes to compare the response and remission rate against random allocation to treatment as reference. For this purpose, we first generated a reference dataset consisting of 1 million simulated outcomes by randomly assigning the subjects to PE regimes. To estimate the effect of optimized treatment allocation, the response to each intervention separately was calculated for each patient using the metamodels described above, then used the predicted outcome for estimating the MADRS score after treatment. Each patient was then virtually assigned to the treatment predicted to yield the best outcome, regardless of the actual assignment in the clinical trial. The response and remission rates were then calculated for each dataset independently.

For the random assignment to treatment, the response rate was 66.7 ± 5.8% (mean ± standard deviation), and the remission rate was 59.9± 5.4% (Fig. 5A). As expected, these values were very close to the observed response and remission rates, 62.1% and 57.6%, respectively (Fig. 5A). When each subject was allocated to the PE regime predicted to yield the best outcome, the response rate was 93.5%, and the remission rate was 87.9% (p<0.05, one sample t-test) (Fig. 5A). This corresponds to an increase in response and remission rates of 30% and 28%, respectively, as compared to random assignment to treatment. For comparison, the simulated outcome in case all subjects were allocated to the same PE regime did not outperform random assignment to treatment (Fig. 5A). The actigraphy data analyzed in this study was recorded on a subset of subjects enrolled in the original study [12]. Therefore, we referred to the original dataset, focusing on treatment as usual (TAU) as comparator, and PE as targeted intervention to which a patient can be assigned based on prior information on effectiveness. In the original dataset, the remission rate for TAU was 37.7% (CI: 32.1% -43.0%), and for PE was 47.0% (CI: 41.5% - 52.5%; effect size *vs.* TAU: + 9.3%). Assuming a similar effect size as in the current sample, the remission rate for PE when each patient is assigned to the treatment predicted to yield best response increased to 75.1% (CI: 70.3% - 79.8%; effect size *vs.* TAU: +37.4%) (See Supplementary Fig. S6). The analysis of differences between observed outcome and best predicted outcome at individual level identified better response to another treatment than originally assigned in 21 out of 66 patients (31.8%; Fig. 5B, bottom right quadrant).

**Figure 5.**
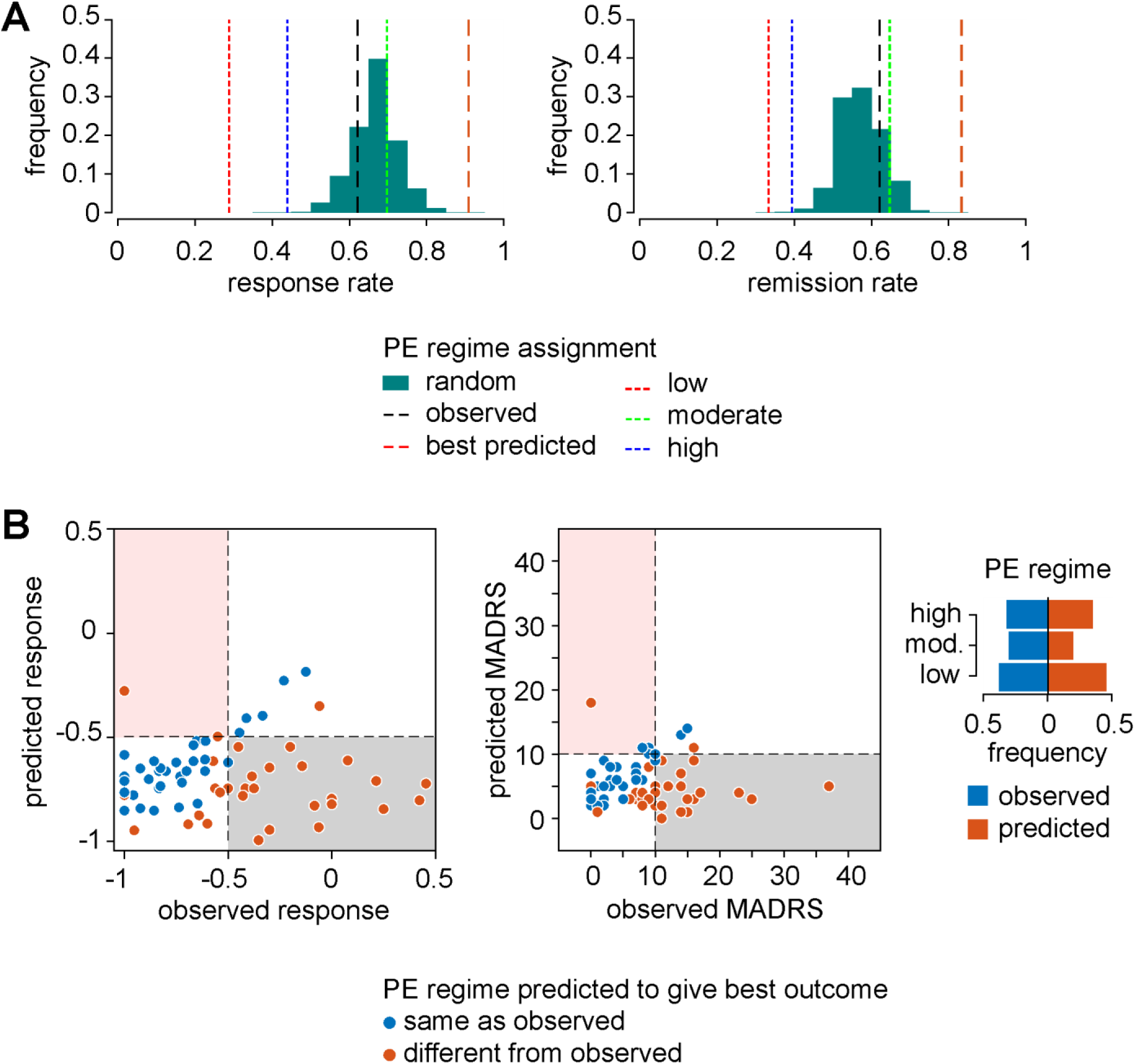
Predicted improvement in patient outcome if *a priori* information on best individual treatment is used for selection of treatment. (A) Counterfactual analysis for predicted response and remission rates using random assignment to treatment (1 million simulations) as comparator. The observed response and remission rates (used for ensemble training) were within 1 standard deviation from the simulated mean rates. Assigning each patient to the PE regime predicted to give best response yield significantly higher response and remission rates (beyond +4 standard deviations; p<0.05, single sample t-test). If all subjects are assigned to the same PE regime, the response and remission rates do not outperform random assignment. (B) Illustration of differences between observed and best predicted response. The grey shaded rectangles highlight the patients with significant improvement in treatment outcome, namely patients predicted to have reached remission if they followed another intervention than originally assigned. Red shaded panel highlights failed predictions, *i.e.*, patients reaching remission in the clinical trial, but predicted not to reach remission. Right panel: distribution of assignment to different PE regimes.

## Discussion

Physical activity, particularly as exercise, produces antidepressant effects, and several biological and psychosocial mechanisms have been suggested to be involved (reviewed in [30,31]). In addition, several features of the PE regime have been shown to impact the outcome (*e.g.*, how structured the PE regime is, as well as session duration, frequency, and intensity), and the optimal protocol varies widely among patients. In our retrospective, explanatory regression analysis, the response to specific PE regimes correlates with distinct subsets of features describing the patient’s patterns of activity. While it is not possible to claim individual protocol optimization, our results suggest it may be possible to identify a PE regime to yield the best results for an individual patient. Using a prospective, predictive approach, we estimated the potential improvement in patient outcome if a priori information on response to different treatment options is used instead of random assignment. The simulation predicted improved response in about 30% of the patients as compared to random assignment to treatment.

Ensemble learning has been developed to improve predictive accuracy by combining the predictive power of multiple models, and it outperforms approaches based on model selection because it does not assume the underlying data-generating model is included in the model list. The design of analyses had to accommodate the asymmetry of the feature matrix (low number of patients per group, high number of features). The diversity of models, combined with the Bayesian model averaging, is supposed to better account for the heterogeneity within the group of patients suffering from major depressive disorder. The ensemble pruning criteria were arbitrarily generous in order to both capture optimum accuracy and accommodate some degree of gradual degradation of performance. The rationale is that the metamodel is implicitly overfitting, since model selection criteria capitalize on fitting accuracy in the training dataset. However, the generalizability of the metamodel presumably increases as less accurate models are included. The Bayesian analysis of coefficients highlights the global differences among ensembles modelling the response to treatment. The features used for modelling (except age) can be assigned to 3 main categories, namely descriptors of circadian profile, circadian entrainment, and ultradian patterns. With the notable exception of propensity to sustain activity (which is a hybrid feature combining distribution with sequence analysis), enriched features belong to descriptors of circadian profile and circadian entrainment, suggesting that ultradian patterns do not have a major contribution to predicting the response to PE as antidepressive intervention. Weaker circadian entrainment, in general, predicts better response to low intensity PE, but worse response to moderate and high intensity PE. We have applied a similar data analysis pipeline for the modelling the response to iCBT or ketamine [15]. For iCBT, weaker circadian entrainment correlated with better response, similar to low intensity PE. In contrast to iCBT and PE, the most relevant correlates of the response to ketamine were descriptors of circadian profile and ultradian patterns of activity [15]. This further emphasizes the differences in feature subsets correlating with the response to different antidepressive therapies.

The patient groups we have analysed were homogenous in terms of inclusion criteria and data collection (including recording conditions, device, and compliance). This allowed a counterfactual evaluation of the benefit of using *a priori* information of the response to treatment as compared to random assignment. The precision of aggregated prediction corresponds to an average error between 2 and 4 points on MADRS scale after treatment for an initial MADRS score between 20 and 30. However, we can identify several caveats for this investigation. First, the accuracy of aggregated predictions has not been verified on external datasets, and there is a risk of overfitting the training dataset. Second, there is no consensus for how to balance accuracy and predictive power for linear regression ensembles, which means we currently do not have definite criteria to drive the pruning of the ensembles. In our datasets, we observed very small differences between predicted outcomes when varying the ensemble size used. Third, the use of hard thresholds for patient stratification, such as for defining remission (MADRS_post_<10) or responder status (response<-0.5), as well as “treatment option with best predicted outcome”, may yield unrealistic estimates. This implies that relevance of deviation from true (observed) response to treatment and absolute MADRS score varies across the range: a predicted response between -1 and -0.5 means “significant improvement”, and a predicted MADRS <10 (remission) is the desirable outcome of any given treatment. Visual inspection of the data yielded two remarkable subgroups. The first group consisted of patients for which most likely an alternative treatment option would yield better outcome, containing about one third of the patient group. The second group consisted of patients for which we could not identify a treatment option that would significantly decrease symptom severity (*i.e.*, classified as responder or in remission). The former illustrates the gain over random assignment (*i.e.*, without prior indication of efficacy as criterion for assignment to treatment), the latter corroborates the contribution of prior indication of treatment efficacy, namely that an alternative treatment should be considered (*i.e.*, outside the range of options we could test, such as CBT, antidepressants, or combinations thereof).

The diversity of protocols used in randomized clinical trials provides little support for estimating the success rate for standardized regimes, but recent meta-analyses found PE to be effective as antidepressive intervention, with remission rates comparable to pharmacotherapy [9,32]. In our cohort, the remission rate following random assignment to treatment was 57.1% and was estimated to increase by 28% if prior information on intervention efficacy is used for treatment assignment. Most randomized clinical studies include a comparator group of usual care or TAU, which is typically defined as treatment according to national guidelines followed up by general practitioners ( *i.e.*, no particular focus on individual follow-up or personalized treatment). In primary care, the remission rate has been estimated to be between 26% and 35% for TAU [33,34]. The success rate increases by 30-35% when using enhanced evidence-based care (EEC; measurement-based care and algorithm-guided therapy) [35]. Targeted interventions in primary care reach remission rates around 54% for monotherapy (antidepressants or psychotherapy) and 67% for combined antidepressants and psychotherapy (∼24% increaseover monotherapy) [33]. Similarly, a recent re-analysis of STAR*D data has shown that sequential antidepressant interventions including switching and augmentation yielded a combined increase of remission rate of ∼37% from first step (citalopram) [4]. The improvement in treatment outcome in our prospective counterfactual analysis does not appear overestimated, and the percentage of patients predicted to have a better treatment outcome is in line with previously reported increase in remission rate following stepwise, measurement-based adjustments in antidepressant treatment [35].

### Limitations

The data we have analysed is derived from a clinical trial where the participants were closely monitored. The population sample may be biased because (1) there are no data available on the number of patients fulfilling the inclusion criteria which declined to take part in the trial; (2) the probability to drop-out was most likely higher among non-responders, which artificially inflates the remission rate at the end of the 12-week trial (see also [36] for review). The response and remission rates in our dataset were comparable to targeted interventions and EEC [33,35], and the inflation as compared to observational studies can be due to the close follow-up of each subject. Therefore, the percentage of patients for which the predicted outcome was better than random assignment to treatment and the relative change in response and remission rates, are probably more informative than the theoretical estimation of response and remission rates.

When training machine learning models, the limitations of the training dataset are typically carried over in predictive applications. For instance, the range of predicted outcome for an external dataset is projected to a range similar as the training dataset. This can be observed in our prospective application of the ensembles (Supplementary Fig. S4: no predicted worsening following low intensity PE), and is a direct consequence of the selection bias embedded in the training dataset. However, the impact of restricted range of response may not be relevant in prospective applications, since the outcome of treatment is typically evaluated using arbitrary thresholds (responder/non-responder; in remission/not in remission).

The models we have developed for predicting the response to PE as antidepressive treatment yield good performance retrospectively, but implementation in clinical practice must satisfy the following prerequisites: (1) the patient needs to meet the inclusion/exclusion criteria of the original clinical trial [12]; the actigraphy device (preferably similar) must be worn attached to the same location on the patient’s body as in the training dataset (in the hip region). The effects of placement of the recording device on parameters of interest (*e.g.*, features describing sleep, physical activity, or posture) has been discussed extensively [37–39]. In addition, the recording protocol may prevent the calculation of relevant features: in our dataset, for instance, we could not estimate the relative amplitude of circadian rhythms (RA; see [19]), because the recording device was not worn at night. To make best use of the data available, we focused the design of feature extraction pipeline on normalized profiles, distributions, and first order dynamics with the explicit aim of capturing device-independent features. The feature selection procedure was data-driven, and we cannot exclude that alternative feature subspaces would be relevant for different applications (*e.g.*, differential diagnosis for psychiatric conditions with overlapping symptoms). Therefore, we believe that our data analysis pipeline provides a sound basis for further applications based on activity monitoring as objective measurement. While we provide proof-of-concept for prediction of response to treatment (see also [15,16]) training ML models to support personalized treatment for depression requires standardized actigraphy recording procedure and antidepressive interventions.

### Conclusion

Currently, most patients with mild to moderate depression are treated with antidepressants or psychotherapy, or combinations thereof. There is evidence that physical exercise as alternative treatment can have a positive effect. However, we do not know which patients that will benefit from this treatment or what intensity of activity that is required. The result from the current study suggests that the analysis of patients’ own pattern of activity could assist in choosing antidepressive treatment.

## Data Availability

Data cannot be shared publicly because of the sensitive nature of medical classification.

https://github.com/stefanspulber/Regassa

## Acknowledgements

The collection of the original data was supported by The Vårdal Foundation (RS2009/27), The Swedish Brain Foundation (Hjärnfonden), and the six Swedish counties and regions involved in the study (YF). Data analysis was supported by Swedish Research Council (2019-01191), The Swedish Brain Foundation (Hjärnfonden, FO2016-0116), Torsten Söderberg Foundation (M59/16), and Karolinska Institutet research grants (SC). The funding agencies and sponsors did not have any influence on the conceptualization, design, data collection, analyses, decision to publish, or preparation of the manuscript.

## Author contribution

Conceptualization: SS. Data curation: SS, YF. Formal Analysis: SS, YF. Funding acquisition: YF, SC. Investigation: YF. Methodology: YF, SS. Project administration: SC, YF. Resources SC, YF. Software: SS. Supervision: SC, YF. Validation: SC, YF. SS. Visualization: SS. Writing – original draft: SS, YF. Writing – review & editing: SS, SC, YF.

## Software and data availability

The codes used for data analysis and visualization are available in a public GitHub repository (https://github.com/stefanspulber/Regassa). The actigraphy data can be made available upon reasonable request.

## Bibliography

1. World Health Organization. Depression and Other Common Mental Disorders Global Health Estimates. Global Health Estimates. Geneva; 2017.

2. Verduijn J, Verhoeven JE, Milaneschi Y, Schoevers RA, van Hemert AM, Beekman ATF, et al. Reconsidering the prognosis of major depressive disorder across diagnostic boundaries: full recovery is the exception rather than the rule. BMC Med. 2017;15: 215. doi:10.1186/s12916-017-0972-8

3. Sansone RA, Sansone LA. Antidepressant adherence: are patients taking their medications? Innov Clin Neurosci. 2012;9: 41–6.

4. Pigott HE, Kim T, Xu C, Kirsch I, Amsterdam J. What are the treatment remission, response and extent of improvement rates after up to four trials of antidepressant therapies in real-world depressed patients? A reanalysis of the STAR*D study’s patient-level data with fidelity to the original research protocol. BMJ Open. 2023;13: 63095. doi:10.1136/bmjopen-2022-063095

5. Casacalenda N, Perry JC, Looper K. Remission in major depressive disorder: a comparison of pharmacotherapy, psychotherapy, and control conditions. American Journal of Psychiatry. 2002;159: 1354–1360. doi:10.1176/appi.ajp.159.8.1354

6. Thornicroft G, Chatterji S, Evans-Lacko S, Gruber M, Sampson N, Aguilar-Gaxiola S, et al. Undertreatment of people with major depressive disorder in 21 countries. British Journal of Psychiatry. 2017;210: 119–124. doi:10.1192/bjp.bp.116.188078

7. Morres ID, Hatzigeorgiadis A, Stathi A, Comoutos N, Arpin-Cribbie C, Krommidas C, et al. Aerobic exercise for adult patients with major depressive disorder in mental health services: A systematic review and meta-analysis. Depress Anxiety. 2019;36: 39–53. doi:10.1002/da.22842

8. Krogh J, Hjorthøj C, Speyer H, Gluud C, Nordentoft M. Exercise for patients with major depression: a systematic review with meta-analysis and trial sequential analysis. BMJ Open. 2017;7: e014820. doi:10.1136/bmjopen-2016-014820

9. Heissel A, Heinen D, Brokmeier LL, Skarabis N, Kangas M, Vancampfort D, et al. Exercise as medicine for depressive symptoms? A systematic review and meta-analysis with meta-regression. Br J Sports Med. 2023;57: 1049–1057. doi:10.1136/bjsports-2022-106282

10. Dhar AK, Barton DA. Depression and the Link with Cardiovascular Disease. Front Psychiatry. 2016;7: 1. doi:10.3389/fpsyt.2016.00033

11. Robison JI, Rogers MA. Adherence to Exercise Programmes. Sports Medicine. 1994;17: 39–52. doi:10.2165/00007256-199417010-00004

12. Hallgren M, Kraepelien M, Öjehagen A, Lindefors N, Zeebari Z, Kaldo V, et al. Physical exercise and internet-based cognitive–behavioural therapy in the treatment of depression: Randomised controlled trial. The British Journal of Psychiatry. 2015;207: 227–234. doi:10.1192/BJP.BP.114.160101

13. Montgomery S, Åsberg M. A new depression scale designed to be sensitive to change. Br J Psychiatry. 1979;134: 382–389. doi:10.1192/BJP.134.4.382

14. Helgadóttir B, Hallgren M, Ekblom Ö, Forsell Y. Training fast or slow? Exercise for depression: A randomized controlled trial. Prev Med (Baltim). 2016;91: 123 –131. doi:10.1016/J.YPMED.2016.08.011

15. Spulber S, Elberling F, Ceccatelli S, Gärde M, Tiger M, Lundberg J. Correlations between patterns of activity and the response to treatment yield distinct signatures for different antidepressive treatments. MedRxiv.org. 2023. doi:10.1101/2023.09.29.23294935

16. Spulber S, Elberling F, Svensson J, Tiger M, Ceccatelli S, Lundberg J. Patterns of activity correlate with symptom severity in major depressive disorder patients. Transl Psychiatry. 2022;12: 226. doi:10.1038/s41398-022-01989-9

17. Hu K, Ivanov PCh, Chen Z, Hilton MF, Stanley HE, Shea SA. Non-random fluctuations and multi-scale dynamics regulation of human activity. Neuroscience. 2004;149: 508 –517. doi:10.1016/j.physa.2004.01.042.Non-random

18. Gonçalves BSB, Cavalcanti P, Tavares GR, Campos TF, Araujo JF. Nonparametric methods in actigraphy: An update. Sleep Science. 2014;7: 158–164. doi:10.1016/j.slsci.2014.09.013

19. Lyall LM, Wyse CA, Graham N, Ferguson A, Lyall DM, Cullen B, et al. Association of disrupted circadian rhythmicity with mood disorders, subjective wellbeing, and cognitive function: a cross-sectional study of 91 105 participants from the UK Biobank. Lancet Psychiatry. 2018;5: 507–514. doi:10.1016/S2215-0366(18)30139-1

20. Edgar N, McClung CA. Major depressive disorder: a loss of circadian synchrony? Bioessays. 2013;35: 940–4. doi:10.1002/bies.201300086

21. Saragiotis C. Lomb normalized periodogram. MATLB Central File Exchange; 2021. Available: https://www.mathworks.com/matlabcentral/fileexchange/22215-lomb-normalized-periodogram

22. Sokolove PG, Bushell WN. The chi square periodogram: its utility for analysis of circadian rhythms. J Theor Biol. 1978;72: 131–160. Available: http://view.ncbi.nlm.nih.gov/pubmed/566361

23. Tackenberg MC, Hughey JJ. The risks of using the chi-square periodogram to estimate the period of biological rhythms. PLoS Comput Biol. 2021;17: 1 –16. doi:10.1371/JOURNAL.PCBI.1008567

24. Chapman JJ, Roberts JA, Nguyen VT, Breakspear M. Quantification of free-living activity patterns using accelerometry in adults with mental illness. Sci Rep. 2017;7: 1 –12. doi:10.1038/srep43174

25. Fasmer OB, Hauge E, Berle JØ, Dilsaver S, Oedegaard KJ. Distribution of active and resting periods in the motor activity of patients with depression and schizophrenia. Psychiatry Investig. 2016;13: 112–120. doi:10.4306/pi.2016.13.1.112

26. Gonçalves B, Adamowicz T, Louzada F, Moreno C, Araujo J. A fresh look at the use of nonparametric analysis in actimetry. Sleep Med Rev. 2015;20: 84–91. doi:10.1016/j.smrv.2014.06.002

27. Raftery AE, Madigan D, Hoeting JA. Bayesian Model Aeraging for Linear Regression Models. J Am Stat Assoc. 1997;92: 179–191.

28. Madigan D, Raftery AE. Model Selection and Accounting for Model Uncertainty in Graphical Models Using Occam’s Window. J Am Stat Assoc. 1994;89: 1535 –1546. doi:10.1080/01621459.1994.10476894

29. Grömping U. Variable importance in regression models. Wiley Interdiscip Rev Comput Stat. 2015;7: 137–152. doi:10.1002/WICS.1346

30. Kandola A, Ashdown-Franks G, Hendrikse J, Sabiston CM, Stubbs B. Physical activity and depression: Towards understanding the antidepressant mechanisms of physical activity. Neurosci Biobehav Rev. 2019;107: 525–539. doi:10.1016/J.NEUBIOREV.2019.09.040

31. Xie Y, Wu Z, Sun L, Zhou L, Wang G, Xiao L, et al. The Effects and Mechanisms of Exercise on the Treatment of Depression. Front Psychiatry. 2021;12: 705559. doi:10.3389/FPSYT.2021.705559/BIBTEX

32. Blumenthal JA, Rozanski A. Exercise as a therapeutic modality for the prevention and treatment of depression. Prog Cardiovasc Dis. 2023;77: 50–58. doi:10.1016/J.PCAD.2023.02.008

33. Dawson MY, Michalak EE, Waraich P, Anderson JE, Lam RW. Is remission of depressive symptoms in primary care a realistic goal? A meta-analysis. BMC Fam Pract. 2004;5: 1–6. doi:10.1186/1471-2296-5-19/FIGURES/1

34. Kolovos S, van Tulder MW, Cuijpers P, Prigent A, Chevreul K, Riper H, et al. The effect of treatment as usual on major depressive disorder: A meta-analysis. J Affect Disord. 2017;210: 72–81. doi:10.1016/J.JAD.2016.12.013

35. Xiao L, Qi H, Zheng W, Xiang YT, Carmody TJ, Mayes TL, et al. The effectiveness of enhanced evidence-based care for depressive disorders: a meta-analysis of randomized controlled trials. Translational Psychiatry 2021 11:1. 2021;11: 1–8. doi:10.1038/s41398-021-01638-7

36. Ormel J, Hollon SD, Kessler RC, Cuijpers P, Monroe SM. More treatment but no less depression: The treatment-prevalence paradox. Clin Psychol Rev. 2022;91: 102111. doi:10.1016/J.CPR.2021.102111

37. Full KM, Kerr J, Grandner MA, Malhotra A, Moran K, Godoble S, et al. Validation of a physical activity accelerometer device worn on the hip and wrist against polysomnography. Sleep Health. 2018;4: 209–216. doi:10.1016/j.sleh.2017.12.007

38. Driller MW, O’Donnell S, Tavares F. What wrist should you wear your actigraphy device on? Analysis of dominant vs. non-dominant wrist actigraphy for measuring sleep in healthy adults. Sleep Science. 2017;10: 132. doi:10.5935/1984-0063.20170023

39. Zinkhan M, Berger K, Hense S, Nagel M, Obst A, Koch B, et al. Agreement of different methods for assessing sleep characteristics: A comparison of two actigraphs, wrist and hip placement, and self-report with polysomnography. Sleep Med. 2014;15: 1107– 1114. doi:10.1016/j.sleep.2014.04.015

