## Supplementary Table 1 for "Individual patterns of activity predict the response to physical exercise as an intervention in mild to moderate depression"

| Feature calculation | Source code | Domain | Reference | Interpretation |
|---|---|---|---|---|
| **period**=main peak between 22 and 26h in Lomb-Scargle periodogram | Adapted from C Saragiotis, 2008 | EN | [1] | Circadian period; should be ~24h, but varies also depending on the precision of the estimation (oversampling factor) |
| **activity onset;** **actoff** = time of first recorded active minute after 4:00 a.m. | OI | CP | N/A | |
| **activity offset;** **acton** = time of last recorded active minute before midnight | OI | CP | N/A | |
| **activity duration;** $\boldsymbol{acttime = actoff - acton}$ | OI | CP | N/A | |
| **middle of active phase;** $\boldsymbol{actmid = \frac{acton + actoff}{2}}$ | OI | EN | N/A | |
| **M10s**=$\boldsymbol{max(smoooth(\{x_i\}))}$; $x_i$= activity counts in 1 min bins, normalized to daily average; smoothing: sliding 600 samples (5h) gaussian window. | OI | CP | Adapted from [2,3] | Magnitude of main circadian peak, normalized to average activity count. |
| **M10L**=location of M10s (h after midnight) | OI | CP | [3] | Timing of occurrence of main circadian peak of activity, typically in the afternoon. |
| **L5s**=$\boldsymbol{min(smoooth(\{x_i\}))}$; $x_i$=activity counts in 1 min bins, normalized to daily average; smoothing: sliding 300 samples (5h) gaussian window. | OI | CP | Adapted from [2] | Activity during the circadian trough, typically during nighttime rest. * |
| **L5L**=Location of L5s (h after midnight) | OI | CP | N/A | Timing of occurrence of main circadian trough of activity, typically before 6 a.m. * |
| **RA**=$\boldsymbol{\frac{M\,10s-L5s}{M10s+L5s}}$ | OI | CP | [2,4] | Relative amplitude of circadian rhythms. Varies between 0 (constant activity throughout the day) and 1 (no activity whatsoever for at least 5h straight). * |
| **alphaFul; alphaShort; alphaLong** = Scaling exponent from detrended fluctuation analysis (DFA) on log-equally spaced intervals between 4 min and 1448 min (24h 8min); between 4 min | OI | UR | [5,6] | Evaluation of complexity of patterns of activity. [elaborate] |

| | | | | |
|---|---|---|---|---|
| and 215 min (3h 35min); between 256 min (4h 16min) and 1448 min (24h 8min). | | | | |
| **IV**= $\frac{n\sum_2^n (b_i - b_{i-1})^2}{(n-1)\sum_1^n (b_i - \bar{b})^2}$; $b_i$= activity counts binned over 5, 30, or 60 min | OI | UR | [2,7] | Estimates fragmentation of activity. It has been shown that the bin width is relevant for pairwise comparisons between conditions [7], therefore we calculated IV in different bins. In our dataset it appears that IV5 and IV30 are orthogonal, while IV30 and IV60 are highly correlated. |
| **IS**= $\frac{n\sum_1^p (b_h - \bar{b})^2}{p\sum_1^n (b_i - \bar{b})^2}$; $b_i$= activity counts in 5, 30, or 60 min bins; $b_h$=average circadian profile in 5, 30, or 60 min bins; *p*=period (24h) | OI | EN | [2,7] | Estimates synchronization with the light-dark cycle, *i.e.*, circadian entrainment. We applied the same reasoning as above, but it appears that IS5, IS30 and IS60 are highly correlated. |
| **rmssd**= $\frac{1}{N-1}\sum_{j=2}^{N}\sqrt{\frac{1}{p}\sum_{i=1}^{p}(x_{i,j} - x_{i,j-1})^2}$; N= number of days; p=period (1440 samples); $x_{i,j}$= activity counts in the *i*-th bin of *j*-th day, normalized to average activity level on *j*-th day. | OI | EN | N/A | RMS difference between normalized circadian profiles of consecutive days (sequential differences). |
| **rmsep**= $\frac{1}{N}\sum_{j=1}^{N}\sqrt{\frac{1}{p}\sum_{i=1}^{p}(x_{i,j} - \bar{x}_i)^2}$; N= number of days; p=period (1440 samples); $x_{i,j}$= activity counts in the *i*-th bin of *j*-th day, normalized to average activity level on *j*-th day. | OI | EN | N/A | RMS difference between normalized individual days and average normalized circadian profile (deviations from average profile). |
| **ddv**= $\sqrt{\frac{\sum_{j=2}^{N}\sum_{i=1}^{p}(x_{i,j} - x_{i,j-1})^2}{\sum_{j=1}^{N}\sum_{i=1}^{p}(x_{i,j} - \bar{x}_i)^2}}$; N= number of days; p=period (1440 samples); $x_{i,j}$= activity counts in the *i*-th bin of *j*-th day, normalized to average activity level on *j*-th day. | OI | EN | N/A | Day-to-day variability estimated as the root ratio between squared sequential differences and the squared deviations from average profile. It cannot be calculated on individual days. For random samples, ddv is approximately $\sqrt{2}$. ddv decreases with consistent circadian profile and small differences between consecutive days. |
| $P(x_{i+1} \geq x_i \mid x_i \in b_j) = m * center(b_j) + n$ **pHiSlope**= $m$; **pHiIcept**= $n$ $\{b_j\}$= distribution of activity in log-equally spaced bins. | OI | UR | N/A | Propensity to sustain activity, estimated as the slope of probability to maintain or increase activity in the next minute calculated for activity levels in log-equally spaced bins. The intercept can be interpreted as the likelihood to increase activity from 1 activity count/min, but has little biological relevance given that the distribution is truncated to remove very low activity levels associated with resting/sleep. In |

addition, the regression line will always approach 0 towards the right tail of the distribution, and slope and intercept are highly correlated.

**Abbreviations:** OI – own implementation in Matlab™, adapted from published reports. N/A – not applicable, original contribution. Domain refers to an arbitrary classification of features based on the core aspects they capture by design. Some features can be assigned to more than one domain, but we stick to single domain classification for simplicity. CP – circadian profile. UR – ultradian rhythms. EN – circadian entrainment. Note that the calculation of variability across days for all features is considered to belong to circadian entrainment (EN).

* - Although typically very informative, these features were excluded from further calculations because nighttime activity is not recorded. The actigraph was worn on a belt only during the active phase of the day and subjects were instructed to recharge the recording device overnight.

**References**


1. Saragiotis C. Lomb normalized periodogram. 2021.
2. Gonçalves B, Adamowicz T, Louzada F, Moreno C, Araujo J. A fresh look at the use of nonparametric analysis in actimetry. Sleep Med Rev. 2015;20:84–91.
3. Ekholm B, Spulber S, Adler M. A randomized controlled study of weighted chain blankets for insomnia in psychiatric disorders. Journal of Clinical Sleep Medicine. 2020;16:1567–1577.
4. Lyall LM, Wyse CA, Graham N, Ferguson A, Lyall DM, Cullen B, et al. Association of disrupted circadian rhythmicity with mood disorders, subjective wellbeing, and cognitive function: a cross-sectional study of 91 105 participants from the UK Biobank. Lancet Psychiatry. 2018;5:507–514.
5. Hu K, Ivanov P, Chen Z, Carpena P, Eugene Stanley H. Effect of trends on detrended fluctuation analysis. Phys Rev E. 2001;64.
6. Spulber S, Conti M, Elberling F, Raciti M, Borroto-Escuela DO, Fuxe K, et al. Desipramine restores the alterations in circadian entrainment induced by prenatal exposure to glucocorticoids. Transl Psychiatry. 2019;9:263.
7. Gonçalves BSB, Cavalcanti P, Tavares GR, Campos TF, Araujo JF. Nonparametric methods in actigraphy: An update. Sleep Science. 2014;7:158–164.
