## Supplementary Information for "Individual patterns of activity predict the response to physical exercise as an intervention in mild to moderate depression"

**Supplementary Material**

Calculation of prior inclusion probability for a specific feature $X_i$

$$\Pr(X_i) = \frac{N\ models\ including\ X_i}{N\ models\ possible} \quad (1)$$

For a given complexity level, the total number of models possible to train is equal to the binomial coefficient $\binom{n}{k}$, and the number of models including a specific feature is the binomial coefficient $\binom{n-1}{k-1}$, where n = total number of features available, and k = model complexity (number of independent variables in the model). Therefore equation (1) can be rewritten as follows:

$$\Pr(X_i) = \frac{\sum_{k=0}^{5}\binom{n-1}{k}}{\sum_{k=0}^{6}\binom{n}{k}} \quad (2)$$

Solving (2) numerically for n=26 yields $\Pr(X_i) = \frac{68\,401}{313\,912} = 0.2179$. The prior inclusion probability of 0.2179 applies to all features, because we trained models for all possible combinations of up to 6 features/model. After pruning the model ensembles, enriched features must have PIP>0.2179, while depleted features have PIP<0.2179.

**Supplementary Fig. S1 Feature set selection**

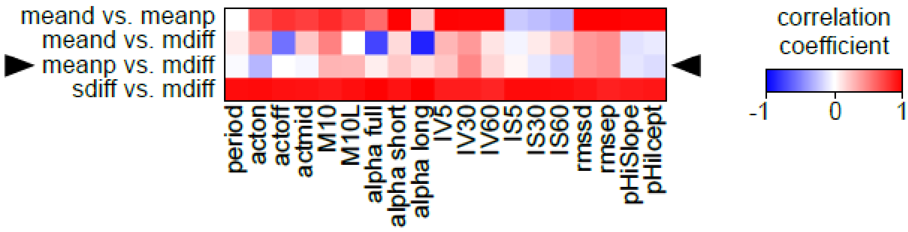


Correlations between main set of features calculated as daily average (meand), or estimated on full-length recording or average circadian profile (meanp), sequential differences in daily values (sdiff), and variance of daily values (*i.e.*, sequence is ignored; mdiff). The lowest correlation coefficients are observed between meanp and mdiff (arrowheads), suggesting that the information carried by the two feature sets is complementary (non-overlapping).

**Supplementary Fig. S2 Feature selection**

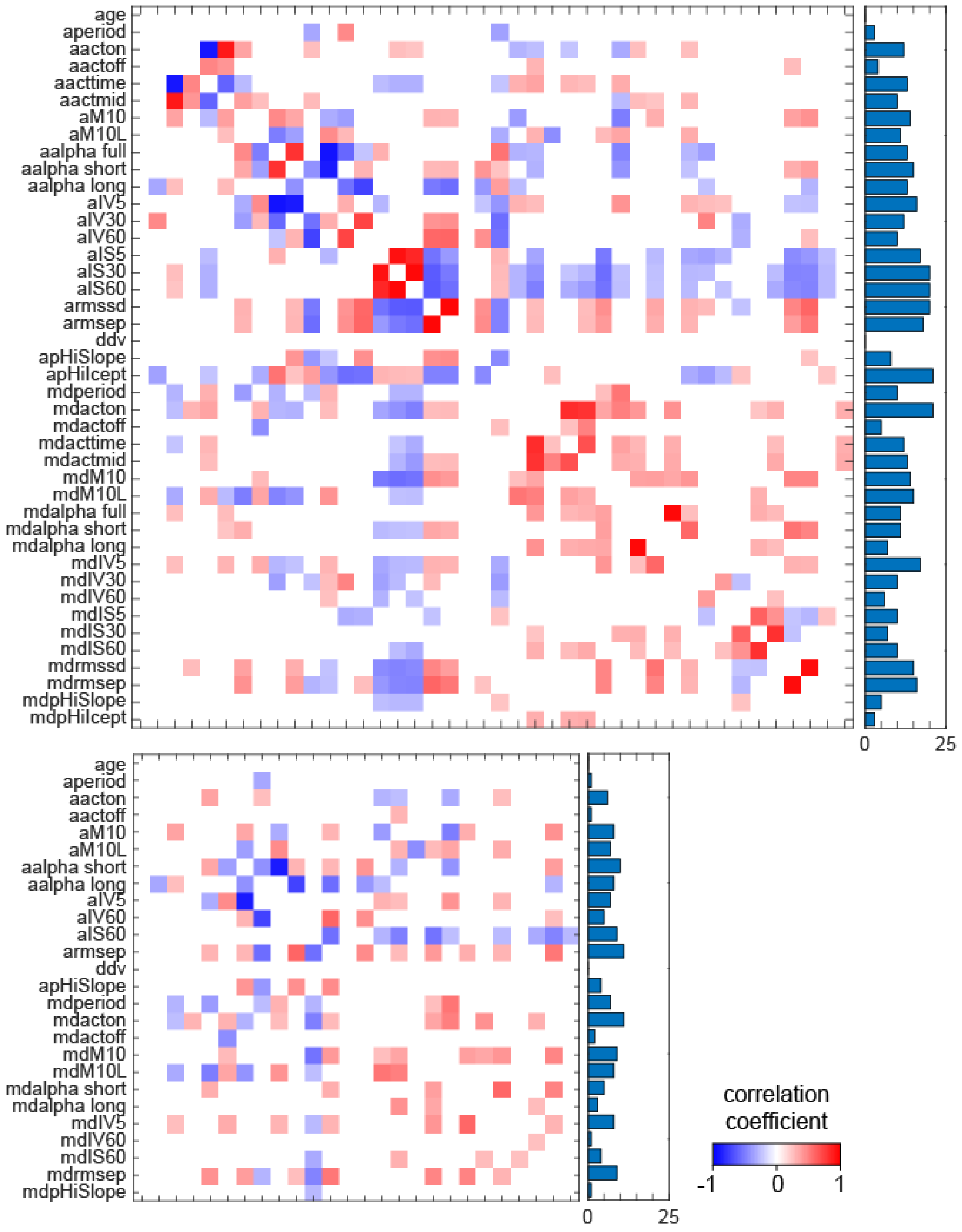


Top panel: pair-wise correlations of all features included in the two feature sets (FDR correction). The clusters of highly correlated and conceptually related features were pruned to include preferentially the features with least correlations in the cluster. Bottom panels: pair-wise correlations after feature selection.

**Supplementary Fig. S3 Trained ensembles**

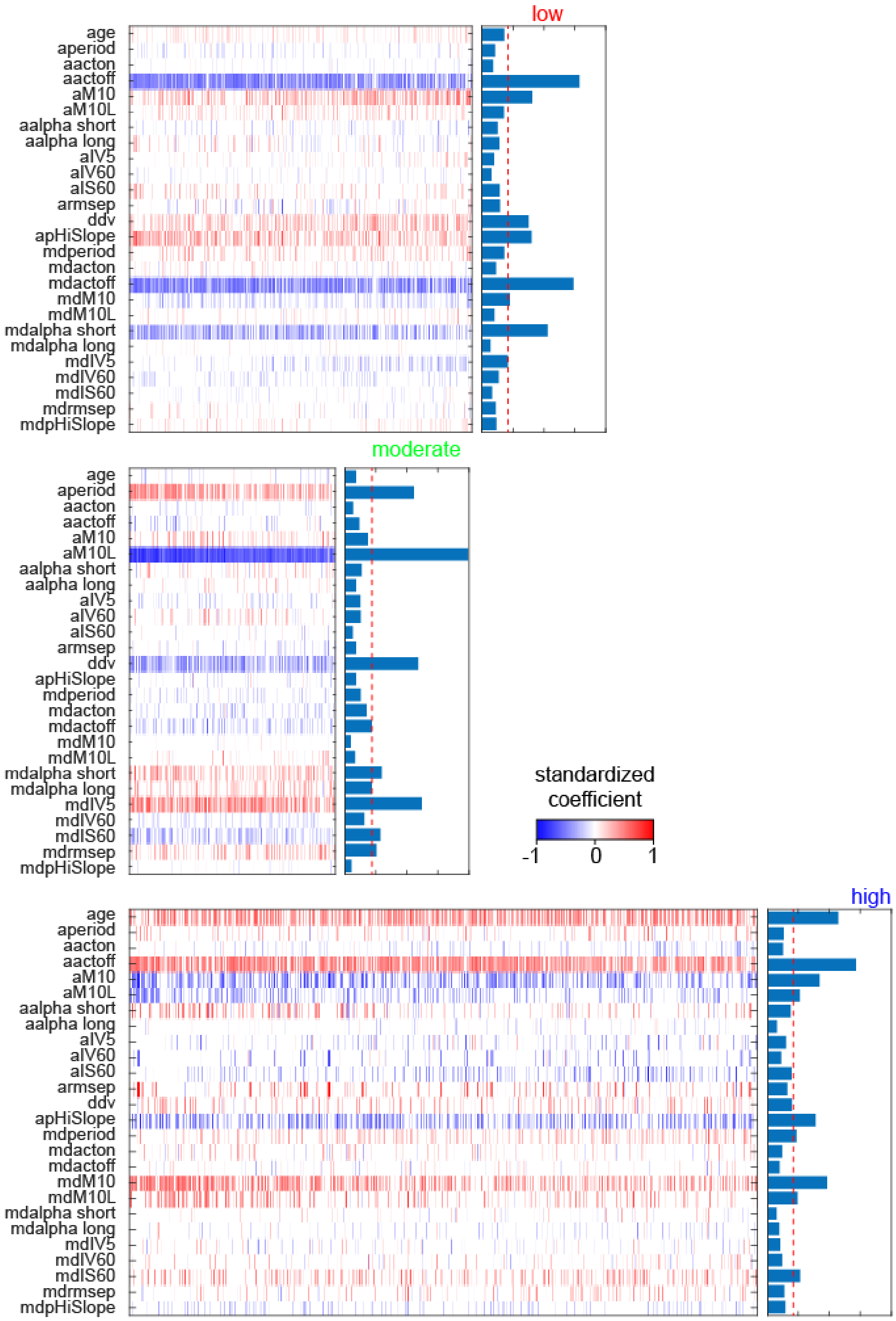

**Supplementary Fig. S4 Metamodel performance analysis.**

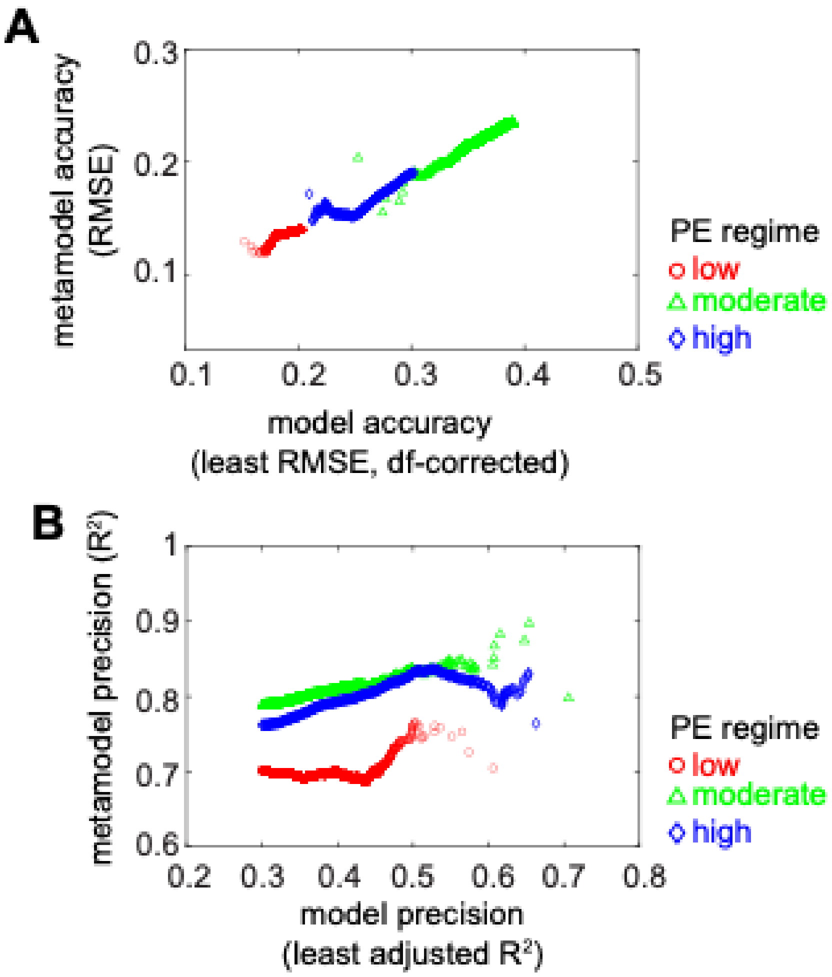


Illustration of metamodel performance in a cumulative fashion: (A) accuracy measured as RMSE vs. observed values; (B) precision estimated by the coefficient of determination, $R^2$. Note that the horizontal axis depicts the accuracy (A) and the precision (B) of the model with worst performance included in each ensemble (see Fig. 5B and the main text for a detailed description of the metamodel). The accuracy of the metamodel prediction increases in the beginning, but then degrades progressively as individual models with decreasing accuracy are added to each ensemble. In contrast, the precision of metamodel prediction does not degrade as the size of the ensembles increases and is consistently higher than 0.7.

**Supplementary Fig. S5 Comparison between predicted outcomes for all PE regimes and subjects.**

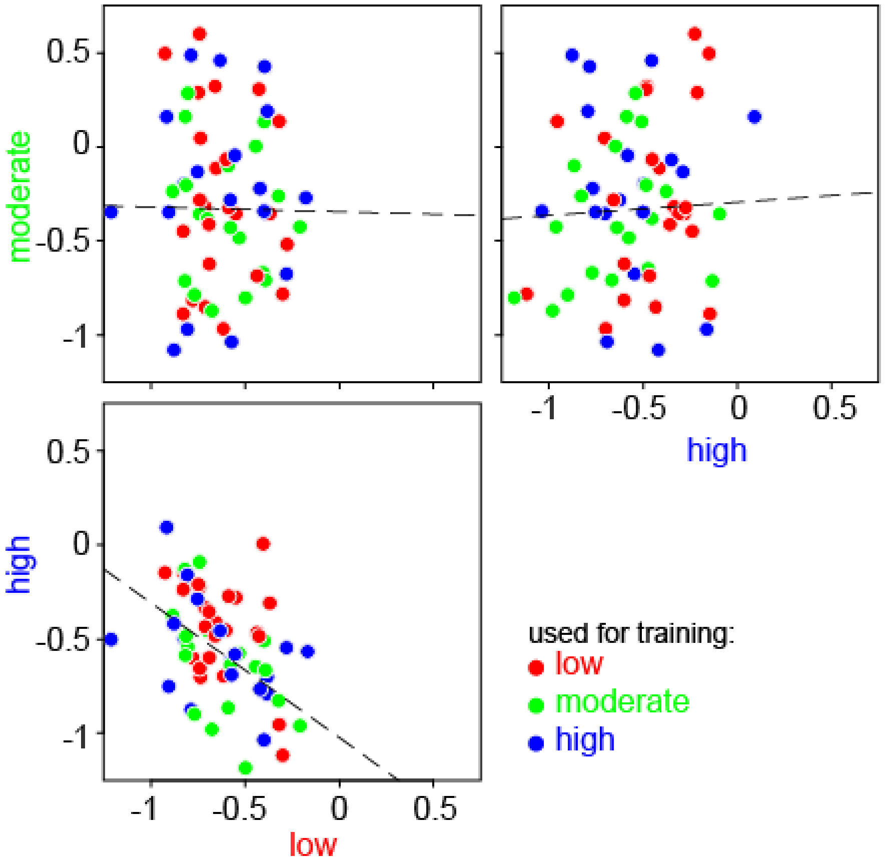


Dashed lines indicate regression slopes estimated using the Theil-Sen method (for illustration purposes only). Note the predicted response to moderate intensity PE is independent for the predicted response to low or high intensity PE, while the response to low and high intensity PE are largely inversely correlated.

**Supplementary Fig. S5 Predicted improvement in treatment outcome provided each patient is assigned to the PE regime predicted to yield the best response**

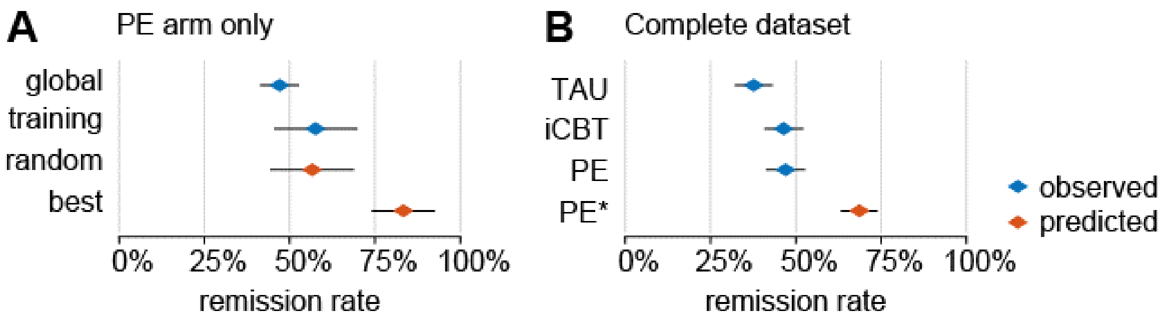


(A) Evaluation of potential selection bias and estimated improvement. The remission rate was slightly higher in the subset of patients used for model training as compared to the global population assigned to PE as antidepressive intervention. The effect size was estimated as (best-random)/random and was applied on the complete dataset in (B). (B) Illustration of improvement in treatment outcome provided each patient is assigned to the PE regime predicted to yield best response. The relative increase in remission rate between PE* and PE is the effect size calculated in (A). Note the net departure from remission rate for TAU as compared to observed remission rate following random allocation to treatment arms in the original study.
